# Preeclampsia-Associated Microbiome Alterations Across Maternal Niches and Infant Gut

**DOI:** 10.1101/2025.08.25.25334409

**Authors:** Courtney L. Birchall, Daniela Vargas-Robles, Dylan C. Hickey, Rose Cohen-Sandler, Monalisa Dmello, Heidi K. Leftwich, Ana Maldonado-Contreras

## Abstract

**Background:** The maternal microbiome has been implicated in the development of preeclampsia. Here, we comprehensively define the oral, vaginal, and gut microbiomes of women with preeclampsia and their infants and compare them with normotensive controls in the US.

**Methods:** Women with and without preeclampsia were recruited during delivery. Oral, vaginal, and gut samples were collected before rupture of membranes in women planning a vaginal delivery. Infant stool samples were collected during the first week of life. Microbiome profiling was performed using *16S rRNA* gene sequencing for oral, vaginal, and infant stool and shotgun metagenomic sequencing for maternal stool. We performed microbiome analyses comparing groups with and without preeclampsia and those with and without severe features of preeclampsia.

**Results:** We recruited 49 and 51 women with and without preeclampsia, respectively. Women with preeclampsia exhibited lower oral microbial richness, distinct composition, and greater interindividual variability than controls. Higher oral microbiome diversity during delivery correlated with lower blood pressure postpartum, regardless of preeclampsia status. Infants of mothers with severe preeclampsia were depleted of gut *Bifidobacterium*. Other differences in the microbiomes emerged after stratification by ethnicity. Hispanic women with preeclampsia exhibited reduced oral richness and shifts in oral nitrate-reducing taxa; reduced vaginal richness and decreased abundance of vaginal *Lactobacillus jensenii;* and depletion of Clostridia species and *Akkermansia* in the gut.

**Conclusions:** Preeclampsia was associated with alterations in the maternal oral, vaginal, and gut microbiomes, depending on the mother’s ethnicity. Infants born to women with preeclampsia exhibit a reduction in the abundance of beneficial *Bifidobacterium*.

## Introduction

Preeclampsia, a hypertensive disorder of pregnancy, is a leading cause of maternal and neonatal morbidity and mortality worldwide^1^. In the United States (US), preeclampsia affects up to 10% of pregnancies, and its incidence continues rising^2^. To prevent preeclampsia, the current standard of care is the daily ingestion of low-dose aspirin, which only reduces the risk by approximately 20%^3,4^. The only curative treatment for preeclampsia remains delivery, which frequently occurs preterm to prevent disease progression and prevent maternal and fetal morbidity and mortality.

Several biological mechanisms and risk factors have been implicated in the development of preeclampsia, including abnormal placentation, endothelial dysfunction, immune dysregulation, genetic predisposition, maternal age, BMI, and the microbiome. The microbiome offers new opportunities to understand the etiology of the disease and to identify potential interventions.

Altered oral, vaginal, and gut microbiota have been associated with preeclampsia^5–9^. Studies in mouse models have shown that microbiome differences in preeclampsia can be causal, influencing blood pressure, placental development, and fetal growth through immune and metabolic pathways^8,10^. Inoculation of mice with gut microbiota from women with preeclampsia induces hypertension, proteinuria, and impaired fetal and placental growth, alongside a shift toward pro-inflammatory immune responses, including a reduction in regulatory T cells (Tregs) — a change also described in pregnant women with preeclampsia and implicated in abnormal placental development^8,11–13^. Short-chain fatty acids (SCFAs), produced by bacterial fermentation of dietary fibers, appear central to these effects. Mice lacking SCFA-producing bacteria or raised germ-free show impaired placental growth and vascularization, whereas SCFA supplementation restores normal placental development^10^. These findings support the hypothesis that depletion of beneficial microbial functions, including SCFA production, may contribute to preeclampsia pathogenesis.

Most work examining the association between the microbiome and preeclampsia has been conducted outside the US and is focused on the gut microbiome, with only a few examining oral and/or vaginal microbiota. These studies do not represent populations in the US, who would have very different environmental factors that directly impact their microbiome, such as diet, lifestyle, and stressors, which could result in different outcome^14–18^. Here, we aimed to determine the oral, vaginal, and gut microbiome dependencies linked to preeclampsia in patients in the US. Moreover, we will investigate whether potential alterations in the maternal microbiome influence the neonatal initial microbiome composition, which is key for immune development and programming^19,20^.

## Methods

### Study Design

We recruited pregnant women with preeclampsia and normotensive controls at the time of delivery at UMass Memorial Health between January 2023 and March 2024. Preeclampsia was defined as new-onset hypertension after 20 weeks’ gestation (≥140 systolic or ≥90 diastolic) along with new-onset proteinuria (≥300 mg proteinuria/24 hours) and/or a severe feature. Severe features of preeclampsia included severe hypertension (≥160 systolic or ≥110 diastolic), impaired liver function (AST and/or ALT ≥2x normal), renal insufficiency (creatinine ≥1.1 or a doubling of baseline), persistent right upper quadrant/epigastric pain, neurologic symptoms, pulmonary edema, HELLP syndrome (<u>h</u>emolysis, <u>e</u>levated liver enzymes, low <u>p</u>latelets), or thrombocytopenia <100,000^21,22^. All women enrolled were planning a vaginal delivery, known to the Group Beta Streptococcus negative, and had intact membranes at the time of specimen collection. Potential recruits were excluded if they took probiotics regularly, had diagnoses of gestational hypertension or chronic hypertension, and/or received antibiotics or had a known intraamniotic infection during their labor before recruitment. Normotensive controls were women who met the inclusion and exclusion criteria outlined above and did not meet the criteria for preeclampsia during pregnancy before delivery. The study protocol was approved by the UMass Chan Institutional Review Board (Docket # H00020140). Written informed consent was obtained from all participants in the study.

### Sample and Clinical Data Collection

Oral, vaginal, and stool samples were collected before delivery. Oral samples were collected by swabbing the tongue for 20-30 seconds, using Omnigene Oral swabs (Cat# OMR-120, DNAGenotek). Vaginal samples were collected by swabbing vaginal walls for 30 s using Omnigene Vaginal swab (Cat# OMR-130, DNAGenotek). Stool samples were collected by swabbing 1–2 inches into the anus for 30 s using Puritan Hydraflock swabs. We also obtained stool samples (from soiled diapers) from infants before hospital discharge (0–6 days after delivery). We obtained detailed clinical data, including demographics and anthropometric information, pregnancy outcomes, postpartum blood pressure, and medications, via a chart review of electronic medical records.

### Sample Processing

We used the Qiagen DNeasy Powersoil Pro Kit (Cat# 47016, Qiagen) to isolate DNA from all the samples collected. The manufacturer’s protocol was followed except both oral and vaginal swabs were pre-treated with proteinase K (Cat# P8107S, New England Biolabs) and incubated for 2 h at 55 °C to breakdown excess mucus; the anal swabs from mothers were submerged in lysis buffer and mechanically disrupted by bead beating using the Omni International Bead Mill Homogenizer; infant stool samples (25-50 mg) were submerged in lysis buffer and heated to 85°C for 10 min, followed by several cycles of bead beating until the sample was completely homogenized.

### Microbiome Profiling

We determined the oral and vaginal microbiomes of mothers by sequencing the V1-V3 *16S rRNA* gene region^23^ and the infant stool microbiome by sequencing the V3-V4 *16S rRNA* gene region ^24^. The microbiome composition of the mothers’ stool samples was determined using shotgun sequencing. All libraries were sequenced using an Illumina NextSeq 2000 instrument and XLeap-SBS chemistry cartridges; we performed 300bp paired-end sequencing (P2 - 600 cycles. Cat# 20100984, Illumina) or 150bp paired-end sequencing (P4 300 cycles; Cat# 20100992, Illumina) for *16S rRNA* and shotgun sequencing, respectively.

### Bioinformatic Processing of Sequences

To perform quality filtering, denoising, and generation of Amplicon Sequence Variants (ASVs), *16S rRNA* sequences were processed using DADA2^25^ in QIIME2^26^. Taxonomic classification of the oral and vaginal samples was performed using a custom GreenGenes-based database combining sequences from GreenGenes, the Human Oral Microbiome Database, and cervicovaginal references from the Human Microbiome Database previously used by others^27^. The resulting feature tables and metadata were imported into R v.4.4.0 ^28^ using the *qiime2R*^29^ package for the downstream analyses. To improve taxonomic resolution, the top 30 most abundant ASVs unclassified at the species level for each sample type were manually annotated using the NCBI BLAST. Taxonomic assignment of the infant stool dataset was performed using the GreenGenes2 database^30^. ASVs classified as archaea, chloroplasts, and mitochondria, unclassified at the order level or higher, or present in only one sample, were removed. ASVs with relative abundance below 0.1% in the oral/vaginal or infant stool dataset were removed. Normalization was performed by rarefying each dataset separately. Oral and vaginal samples were rarefied to 8,634 sequences/sample, and infant stool samples were rarefied to 43,539 sequences/sample. Good’s coverage index was used to evaluate sequencing depth adequacy, with values exceeding 90% and 99% for the oral/vaginal and infant stool datasets, respectively, calculated using the *QsRutils*^31^ package. Microbiome diversity metrics were computed using the *vegan*^32^ and *phyloseq*^33^ functions.

Whole genome sequences were quality-filtered to remove contaminants and human DNA using KneadData (dec_v0.1)^34^. Taxonomic profiling was performed using MetaPhlAn4 (vOct22)^35^ to identify and quantify species-level genomic bins (SGBs). These SGBs were then converted to Genome Taxonomy Database (GTDB) annotations using the sgb_to_gtdb_profile.py script ^36^ from the MetaPhlAn repository. The resulting GTDB-converted profiles were merged to produce relative abundance species-level genomic bin (SGB) tables. Microbial diversity metrics were computed using the calculate_diversity.R script ^37^ from the MetaPhlAn repository. Samples with 100% unclassified taxa were excluded before diversity analysis to ensure valid metric estimations. The SGB table was subsequently imported into R^28^ and merged with the metadata and diversity measures. A phyloseq object was built and filtered, removing SBG with lower than 0.01% abundance and those present in a single sample were removed. All data analyses and visualizations were conducted using the *phyloseq* ^33^ and *tidyverse*^38^ packages.

The raw sequences and their associated metadata have been deposited in PRJNA1306676 (https://dataview.ncbi.nlm.nih.gov/object/PRJNA1306676?reviewer=e80pa9pcmannrthu27lv2tbe on). The complete code used for the bioinformatics and statistical analysis presented in this study is publicly available in the following GitHub repository: https://github.com/UMassDVR/Preeclampsia_microbiome_analyses_scripts.git

### Statistical analyses

Demographic and clinical variable comparisons among groups were assessed using the t-test/Kruskal-Wallis test or Fisher’s exact test for numerical and categorical variables, respectively. Normality was verified using *shapiro.test* function in R.

### Microbiome Diversity Analyses

Microbial diversity was assessed using alpha and beta diversity analyses. All analyses were performed at the ASV level using the rarefied data. All analyses compared preeclampsia status (preeclampsia vs. normotensive controls) and preeclampsia severity, defined as preeclampsia with and without severe features. Differences in alpha diversity were assessed using linear regression models with each alpha diversity metric as the outcome variable. To meet the assumption of normality for linear regression, logarithmic or Box-Cox transformations were applied to the outcomes when necessary. If normalization could not be achieved, a robust linear model (RLM, *rlm*^39^ function) was used to analyze the data. Specifically, RLM was applied to the Simpson index models for oral samples and all alpha diversity metric models for the analysis of infant stool samples. Differences in beta diversity were assessed using Permutational Multivariate Analysis of Variance (PERMANOVA^40^) with the *adonis2*^32^ and beta dispersion test with *betadisper*^32^ functions on both Bray-Curtis and Jaccard distance matrices.

The full model for the analysis of women included ethnicity, antibiotic use during pregnancy, and maternal BMI as initial covariates. The full model for the infants included maternal preeclampsia status or severity, baby weight, mode of delivery, neonatal ICU admission, mother’s ethnicity, and day of stool collection as initial covariates. Both models were refined to include only the relevant variables using the *step* R function in linear models or manually in the case of RLM and PERMANOVA. The identification of specific discriminant microbial taxa was assessed using MaAsLin2^41^ (Multivariable Association with Linear Models) with a previous filtering to include only taxa in ≥ 20% of samples and with ≥ 15% relative abundance. Significant associations were obtained when the nominal p-value was < 0.050 and the false discovery rate (FDR)-adjusted p-value was < 0.250. FDR correction was applied using the Benjamini-Hochberg procedure.

### Prevalence Analyses

Taxa prevalence among the cohort was assessed and used to define the “core taxa”: present in > 40% of women, and “accessory taxa” present in 20–40% of women for each body site, as previously described^42^. Differences in the prevalence of core and accessory taxa were assessed using Fisher’s exact test, and p-values were adjusted to control for multiple comparisons. Significant associations were obtained when nominal p < 0.050 and a false discovery rate (FDR)-adjusted p-value < 0.250. FDR correction was applied using the Benjamini-Hochberg procedure.

### Prevotella:Bacteroides Ratio

The ratio of *Prevotella* to *Bacteroides* (P:B) was calculated using the genus-level relative abundance from the maternal stool samples. Taxa were aggregated at the genus level for analysis. The ratio was log-transformed to improve normality, and a small pseudocount (<1) was added to manage zero values. We compared the P:B ratios between groups using Wilcoxon rank-sum tests (for medians) and Levene’s test (for variance) in R.

### Community State Type (CST) Profiling

Vaginal microbiotas were classified into Community State Type or CST, as described before^43^. CST classification categorizes vaginal microbiomes into five (CST I-V) distinct groups based on the dominant (or lack thereof) *Lactobacilli* species. Each CST, dominated by different *Lactobacillus* species (CST I: *L. crispatus*-dominated, CST II: *L. gasseri*-dominated, CST III: *L. iners*-dominated, CST IV: non-*Lactobacillus* dominance, CST V: *L. jensenii*-dominated), is linked to varying vaginal health, infection risks, and pregnancy outcomes^44^. To compare CST by preeclampsia status and severity, we used Fisher’s Exact Test ^45^ and the pairwise Fisher Exact Test.

## Results

### Description of study population

A total of 100 pregnant women were recruited: 49 diagnosed with preeclampsia and 51 normotensive controls. The mean maternal age was 31.7 years, with a BMI of 30.5 kg/m^2^, and most participants self-identified as non-Hispanic White (59%). The majority of women delivered vaginally (68%) at term (88%), and only one required ICU admission. Compared to controls, women with preeclampsia had higher BMIs and a lower frequency of term delivery. (**Table 1**). Slightly more than half of the infants were female (55.6%). Infants born to mothers with preeclampsia had lower birth weights and were more frequently admitted to the Neonatal ICU than controls (**Table 1**).

**Table 1.** Demographic, anthropometric, and clinical characteristics of the pregnant women with preeclampsia (PE) and normotensive controls, recruited for the study between January 2023 and March 2024 and their infants.

| Variables | Preeclampsia Status |  | P value<sup>&</sup> |
| --- | --- | --- | --- |
|  | Control | PE |  |
| **n** | **51** | **49** |  |
| Mean age (SD) | 31.61 (5.19) | 31.86 (5.53) | 0.817 |
| Race (%) |  |  | 0.822 |
| White | 35 (68.6) | 32 (65.3) |  |
| Other | 9 (17.6) | 12 (24.5) |  |
| Black or African American | 3 (5.9) | 3 (6.1) |  |
| Asian | 3 (5.9) | 1 (2.0) |  |
| Prefer not to answer | 1 (2.0) | 1 (2.0) |  |
| Ethnicity = Hispanic or Latina (%) | 14 (27.5) | 13 (26.5) | 1.000 |
| Mean BMI (SD) | 28.18 (6.63) | 32.76 (8.41) | 0.003* |
| Diabetes = Yes (%) | 7 (13.7) | 9 (18.4) | 0.719 |
| Mean gestational age, weeks (SD) | 39.89 (0.85) | 37.44 (2.07) | <0.001* |
| Term status ($\geq 37$ weeks) = term (%) | 51 (100.0) | 37 (75.5) | 0.001* |
| Delivery mode = Cesarean Delivery (%) | 12 (23.5) | 20 (40.8) | 0.101 |
| Antibiotics during pregnancy = Yes (%) | 4 (7.8) | 6 (12.5) | 0.664 |
| Mother ICU = Yes (%) | 0 (0.0) | 1 (2.1) | 0.992 |
| Infant sex = Male (%) | 22 (43.1) | 23 (46.9) | 0.856 |
| Mean infant weight, grams (SD) | 3503.84 (440.11) | 2932.80 (588.76) | <0.001* |
| Neonatal ICU = Yes (%) | 4 (7.8) | 15 (30.6) | 0.008* |
| & Numerical and categorical variables were compared using t-test/Kruskal-Wallis or Fisher exact test respectively |  |  |  |

Among all women with preeclampsia, 57% had preeclampsia with severe features (PE-SF). Those with severe features had a significantly lower gestational age at delivery than those without severe features (PE-NSF**; Table S1**). Apart from this maternal difference, no significant demographic, anthropometric, or clinical difference was observed between infants born to women with severe versus without severe features (**Table S1**).

### Maternal oral microbiome

We examined differences in the oral microbiomes by preeclampsia status, adjusting for ethnicity, antibiotic use during pregnancy, and BMI. Compared to controls, women with preeclampsia exhibited lower oral microbial richness (p = 0.040, **Fig. 1A, Table S2**), distinct microbiome composition (p = 0.020), and higher interindividual variability (p = 0.042, **Fig. 1B, C,** and **Table S3**). Women with preeclampsia showed a decreased abundance of *Rothia* and an increased abundance of *Neisseria* (p < 0.050, adjusted p < 0.250, **Fig. 1D**) in their oral cavity.

**Figure 1:**
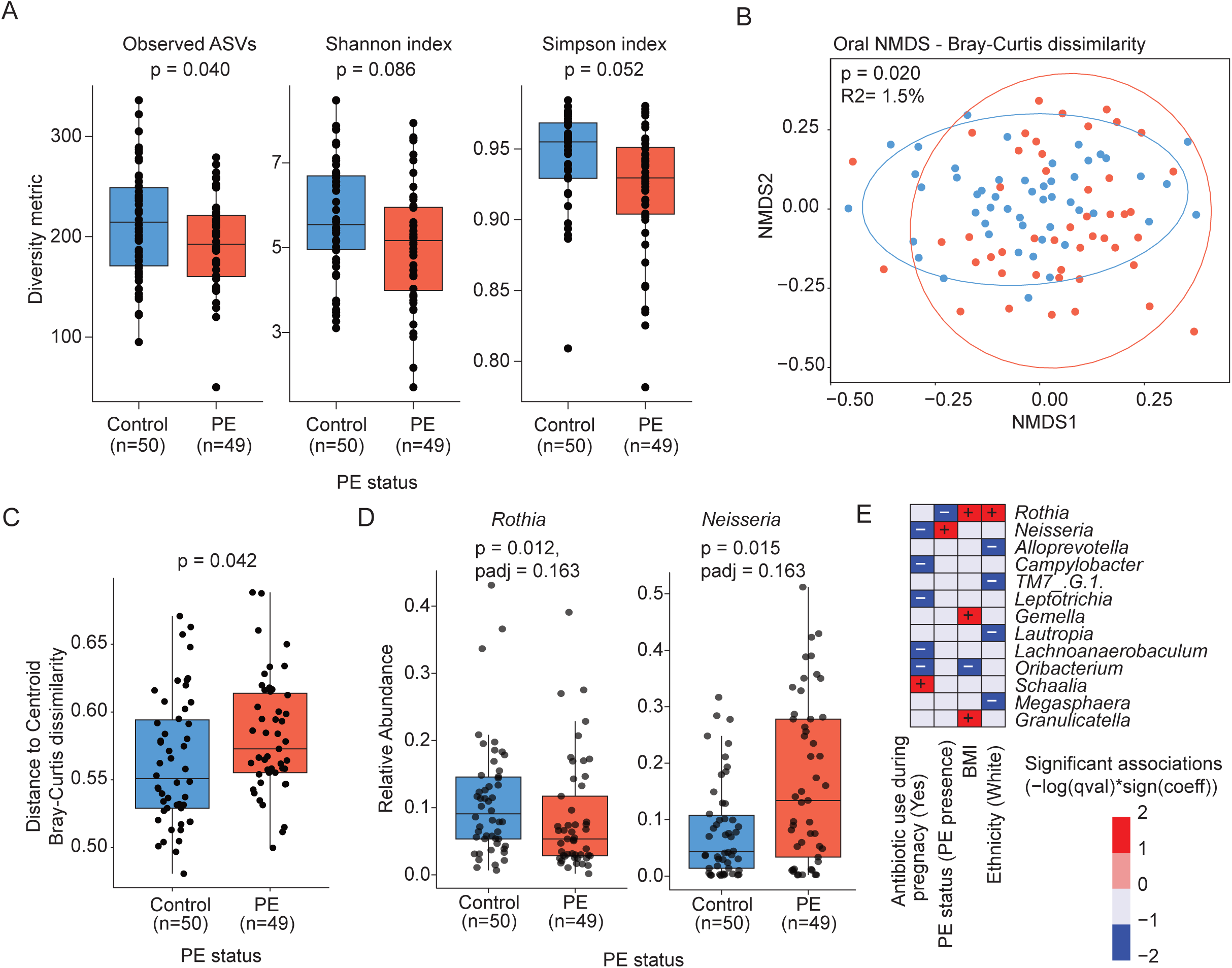
Oral microbiome diversity in pregnant women by preeclampsia (PE) status. (A) Alpha diversity comparisons (Observed ASVs, Shannon, and Simpson indices). P values for alpha diversity analyses were obtained from linear models adjusting by BMI, antibiotic used during pregnancy and ethnicity. (B) Beta diversity analysis using Bray-Curtis distance and non-metric multidimensional scaling (NMDS) ordination, with PERMANOVA results reporting p value and R². The ellipses represent the 95% confidence intervals. (C) Beta dispersion test based on Bray-Curtis distance, comparing distance-to-centroid. (D) Differentially abundant genera (p < 0.050, adjusted p < 0.250, MaAsLin2). (E) Taxonomic heatmap showing associations between bacterial genera and clinical covariates, including preeclampsia status, antibiotic use, BMI, and ethnicity (MaAsLin2).

Both *Rothia* and *Neisseria* are nitrate-reducing genera.

The oral core taxa (taxa at the genus level present in >40% of samples) in women in this cohort showed similar prevalence regardless of preeclampsia status. Among the accessory taxa (taxa at the genus level present in 20–40% of samples), a member of the Bacteroidia class: *Tannerella,* was significantly less prevalent in the preeclampsia group than in the control group (16.3% vs. 40%, p = 0.014, adjusted p = 0.175; **Table S4**). We did not find any differences in oral microbiome alpha or beta diversity between women with preeclampsia, with or without severe features (p > 0.050, **Tables S2 and S3**).

The oral microbiota of women in this cohort was strongly differentiated by ethnicity; Hispanic women exhibited a different microbiome composition compared to non-Hispanic women of all races (p < 0.050, **Fig. S1**). To minimize confounding by ethnicity and better isolate the effects of preeclampsia, we stratified the analysis by ethnic group, focusing on Hispanic or Latina (n=27) and non-Hispanic women who identified as White (n=59; **Table S5**). We will refer to “Hispanic or Latina women” as “Hispanic women” and non-Hispanic women who identified as White as “White women” for the remainder of the manuscript. We excluded women who identified as non-Hispanic Black (n=6), non-Hispanic Asian (n=4), non-Hispanic Other (n=3), or those who selected “Prefer not to answer” (n=1) due to small sample sizes and previously reported differences in the oral microbiome, which could complicate the interpretation of the results^46^. Although ethnicity showed the strongest effect, BMI and antibiotic use also contributed to microbiome diversity and composition (**Tables S2 and S3, Fig. 1E**) and continued being included as covariates in all models.

We observed that White women with preeclampsia had a higher BMI, higher preterm birth rates, and lower gestational age than their control counterparts (p < 0.050; **Table S6**). Hispanic women with preeclampsia had a lower gestational age than their control counterparts (**Table S2**). When comparing White vs. Hispanic women with preeclampsia, we did not observe any differences in demographic and clinical variables (**Table S7**). The oral microbiome of Hispanic women in the preeclampsia group showed higher microbial interindividual variability (p = 0.012, **Fig. 2A**) and reduced relative abundance of *Granulicatella* and its species, *Granulicatella adiacens* (Bacilli class), and nitrate-reducing *Haemophilus parainfluenzae* (Gammaproteobacteria class), compared with Hispanic controls (p < 0.050, adjusted p < 0.250; **Fig. 2B**). Conversely, White women with preeclampsia exhibited a higher relative abundance of the nitrate-reducing *Neisseria* and its species, *Neisseria subflava* (Betaproteobacteria class), and a lower relative abundance of the nitrate-reducing *Rothia* (Actinomycetia class) than White women in the control group (p < 0.050, adjusted p < 0.250, **Fig. 2C**). No differences in the prevalence of core or accessory oral taxa were found between Hispanic and White women (**Table S4**).

**Figure 2.**
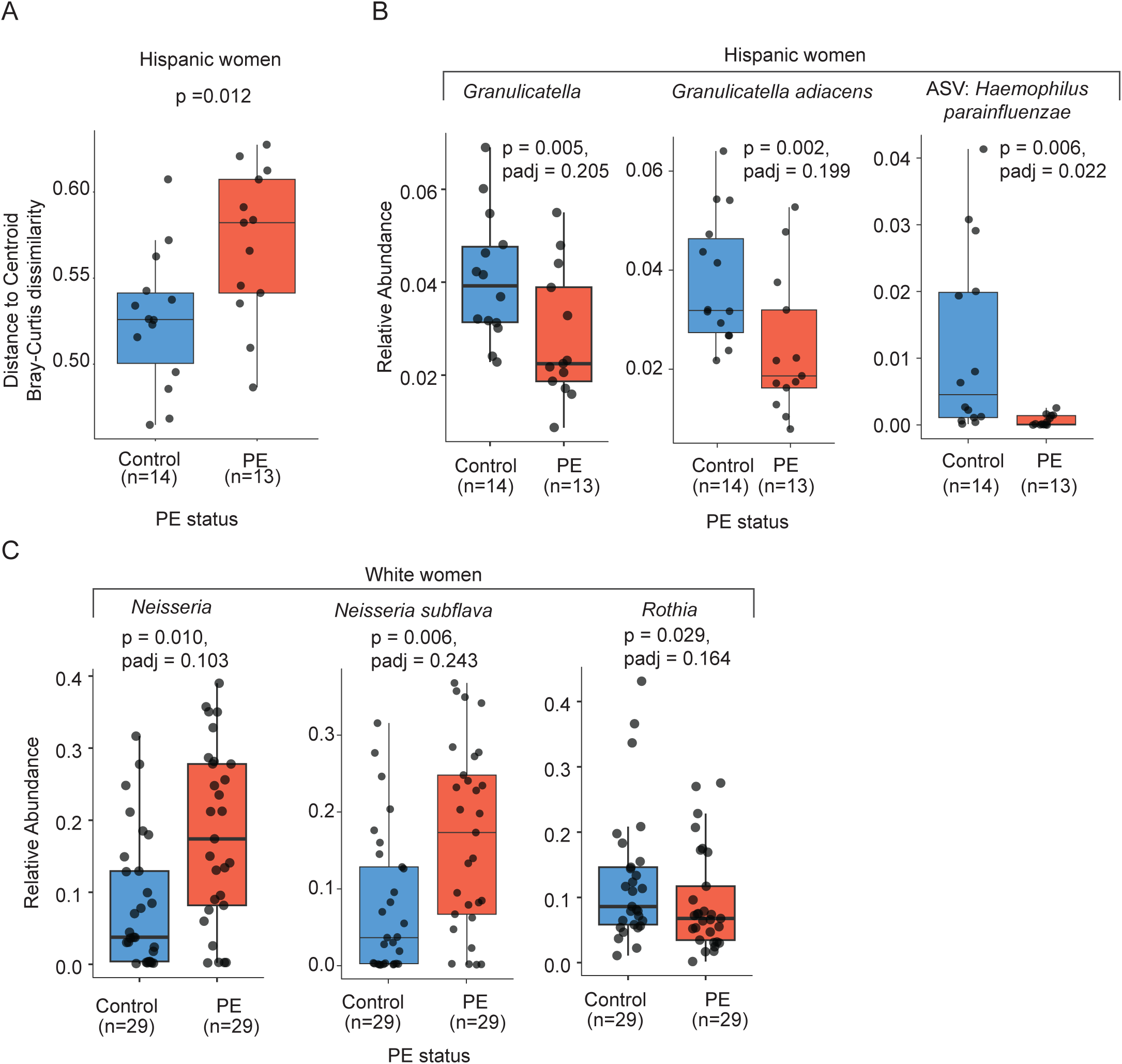
Oral microbiome diversity of pregnant women by preeclampsia (PE) status, stratified by ethnicity (Hispanic or White). (A) Beta dispersion test based on Bray-Curtis distance among Hispanic women, comparing distance to centroid. (B, C) Differentially abundant taxa (p < 0.050, adjusted p < 0.250, MaAsLin2) among Hispanic (B) and White women (C).

In summary, in this US cohort, women with preeclampsia exhibited lower oral microbial richness, distinct microbiome composition, and greater interindividual variability than controls, independent of ethnicity, BMI, and antibiotic use. Preeclampsia was associated with a reduced abundance of *Rothia* and an increased abundance of *Neisseria*, both of which are nitrate-reducing genera. Stratified analyses revealed ethnicity-specific patterns: Hispanic women with preeclampsia showed reduced *Granulicatella* (including *G. adiacens)* and *Haemophilus parainfluenzae*, while White women had elevated *Neisseria* (including *N. subflava*) and reduced *Rothia*. The core oral taxa were consistent across the groups.

### The maternal vaginal microbiome

The vaginal microbiome diversity of women in this cohort did not differ by preeclampsia status (p > 0.050, **Table S2**). However, women with preeclampsia exhibited higher interindividual variability in rare taxa than controls (p = 0.014, **Fig. 3A, Table S3**). The vaginal core taxa showed similar prevalence in women with and without preeclampsia, while the accessory taxa *Enterococcus* (Bacilli class, 30.6% vs 10.0%, p = 0.012, adjusted p = 0.113), *Fusobacterium* (Fusobacteria class, 30.6% vs 12.0%, p = 0.028, adjusted p = 0.113), and *Propionimicrobium (*Actinomycetia class, 32.7% vs 14.0%, p = 0.034, adjusted p = 0.113), were more prevalent in women with preeclampsia than the control group (**Table S4**). No differences were observed in the vaginal microbiome by preeclampsia severity (**Tables S2 and S3**). However, women with severe features of preeclampsia showed a decreased abundance of *Lactobacillus jensenii* compared to those without severe features (p = 0.012, adjusted p = 0.139; **Fig. 2C**).

**Figure 3.**
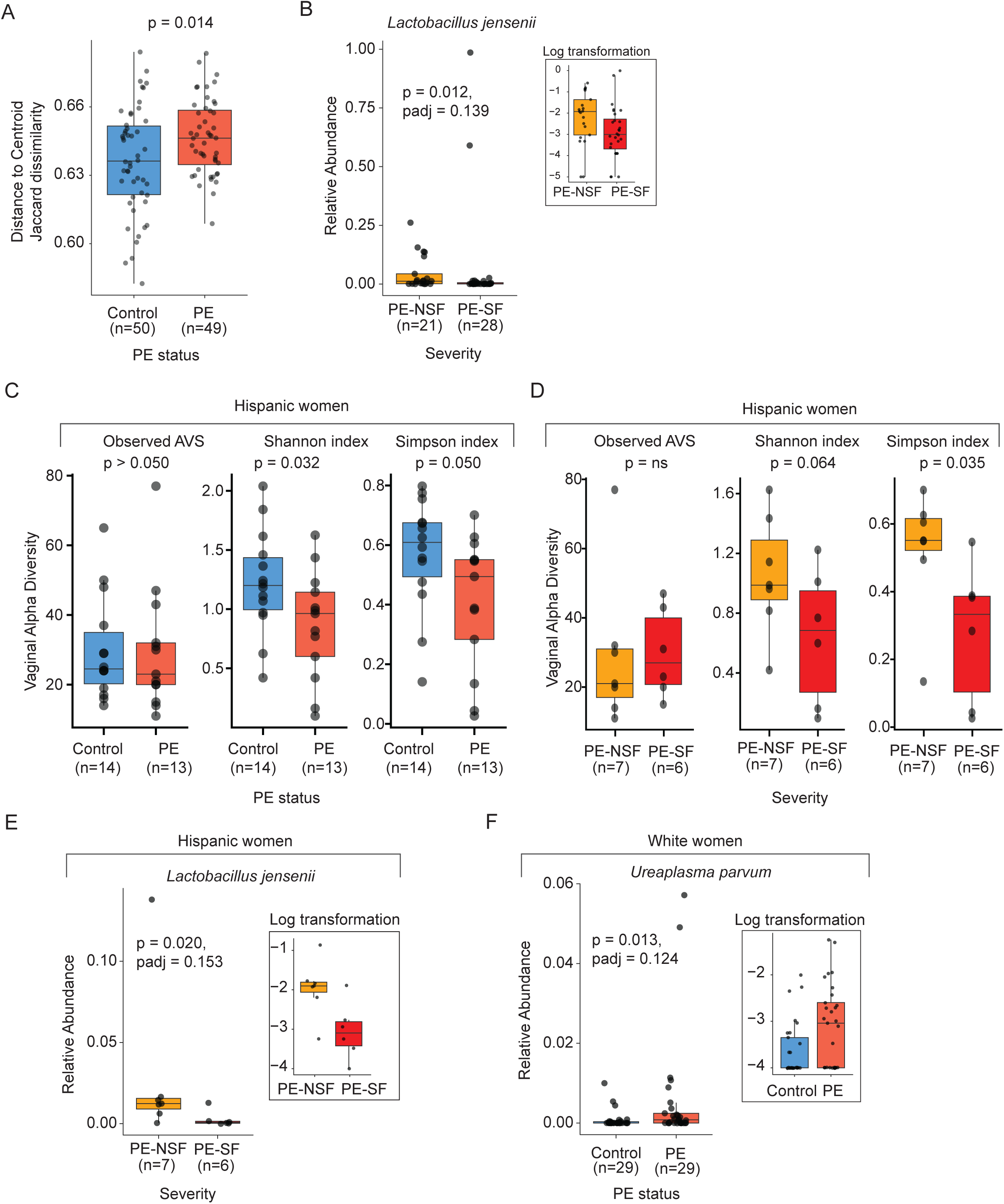
Vaginal microbiome diversity in pregnant women by preeclampsia (PE) status, PE severity, among all women or stratified by ethnicity (Hispanic or White). (A) Beta dispersion test based on Jaccard distance, comparing distance-to-centroid among all women by PE status. (B) Differential abundance of *Lactobacillus jensenii* among all women by PE severity (PE with severe features, PE-SF vs. PE without severe features, PE-NSF; p < 0.050, adjusted p < 0.250, MaAsLin2).(C, D) Alpha diversity comparisons (Observed ASVs, Shannon, and Simpson indices) among Hispanic women by PE status (C) and PE severity (D). P values for alpha diversity analyses were obtained from linear models adjusting by BMI, antibiotic used during pregnancy and ethnicity (when applied). Non-significant p values (p = ns) indicate variables excluded during best-fit model selection. (E, F) Differentially abundant taxa identified by MaAsLin2 (p < 0.050, adjusted p < 0.250) in Hispanic women according to PE severity (E) and White women according to PE status (F).

The vaginal microbiome of women in this cohort also differed by ethnicity (p = 0.029, **Fig. S1**). Therefore, we stratified the participants into two groups—Hispanics and Whites —and compared the vaginal microbiome by preeclampsia status within each group.

Among Hispanic women, those with preeclampsia showed lower alpha diversity (p = 0.032, **Fig. 3A**, **Table S2**) than controls. Moreover, Hispanic women with severe features of preeclampsia had even lower alpha diversity than those without severe features of preeclampsia (p = 0.035, **Fig. 3B**, **Table S2**). Moreover, Hispanic women with severe features of preeclampsia showed a decreased abundance of the beneficial *Lactobacillus jensenii* compared to those without severe features (p = 0.020, adjusted p = 0.153, **Fig. 3C**). No differences in taxa prevalence were observed among Hispanic women by preeclampsia status or severity (**Table S4**).

Among White women, no differences in vaginal microbiome diversity or composition were found by preeclampsia status or severity (**Tables S2 and S3**). White women with preeclampsia exhibited increased abundance of *Ureaplasma parvum (*p = 0.013, adjusted p = 0.124, **Fig. 3D**) and had a higher prevalence of accessory taxa from the Bacilli class, *Enterococcus* (34.5% vs. 6.9%, p = 0.021, adjusted p = 0.246) and *Facklamia* (41.3% vs. 13.8%, p = 0.038, adjusted p = 0.246), than controls (**Table S4**).

BMI was also associated with vaginal microbial diversity in this cohort (**Table S2 and S3**). After stratification by ethnicity, the association between BMI and vaginal microbial diversity remained present only among White women (**Tables S2 and S3**).

We grouped vaginal microbial profiles into Community State Types (CSTs), a classification system developed specifically for the vaginal microbiome based on the dominance (or lack thereof) of specific *Lactobacillus* species^47^. In this cohort, most women had CST I (57%, *L. crispatus-*dominant profile), followed by CST III (29%, *L. iners-*dominant profile), CST II (5%, *L. gasseri-*dominant profile), CST V (4%, *L. jensenii-*dominant profile), and CST IV (4%, non-*Lactobacillus*-dominated profile). No significant differences in prevalence were found in CST by preeclampsia status, including all women or stratified by ethnicity (p > 0.050, **Table S8**). The comparison of CST distribution according to preeclampsia severity was not assessed due to the small sample size among the multiple CST groups, which considerably reduced the power.

In summary, while vaginal microbial diversity did not differ by preeclampsia status, women with preeclampsia showed greater variability in rare taxa and increased prevalence of select accessory taxa. Severe features were associated with decreased *Lactobacillus jensenii.* Ethnicity influenced the vaginal microbiota of women with preeclampsia via reduced diversity and *L. jensenii* abundance among Hispanic women and increased *U. parvum* and accessory taxa among White women. BMI was associated with diversity only among White women. CST distributions did not differ by preeclampsia status or ethnicity.

### Maternal gut microbiome

The gut microbiome diversity of women in this cohort did not differ by preeclampsia status or severity (p > 0.050, **Tables S2 and S3**). However, women with preeclampsia had a lower prevalence of members of the core taxa belonging to the Bacteroidia class, such as *Alistipes* (59.2% vs. 86.3%, p = 0.003, adjusted p = 0.222) and *Bacteroides* (85.7% vs. 100%, p = 0.005, adjusted p = 0.223), than the controls. Among taxa from the Clostridia class, the accessory taxon *Facklamia_A* was more prevalent in women with preeclampsia (40.8% vs. 15.7%, p = 0.007, adjusted p = 0.241), while *Faecousia* was significantly less prevalent (16.3% vs. 51.0%, p = 0.0003, adjusted p = 0.021).

Antibiotic use during pregnancy, reported by 10% of participants, was associated with significant differences in gut microbial alpha diversity (**Table S2**). In contrast to the oral and vaginal microbiomes, the gut microbiome was not significantly associated with ethnicity (**Tables S2 and S3**). However, we observed differences in the gut microbiome according to preeclampsia status when stratifying by ethnicity.

Among Hispanic women, those with preeclampsia presented decreased abundance of Clostridia class (based on taxa aggregated at the class level, p < 0.050, adjusted p < 0.250, **Fig. 4A**) compared to controls. In addition, Hispanic women with preeclampsia showed a lower prevalence of species from the Clostridia and Verrucomicrobiae classes: the core taxon *CAG-170* (7.7% vs. 78.6%, p = 0.0003, adjusted p = 0.030) and the accessory taxon *Akkermansia* (7.7% vs. 64.3%, p = 0.004, adjusted p = 0.257, **Table S4**), respectively. Among White women, those with severe features of preeclampsia had higher interindividual variability than those without severe features (p = 0.043, **Fig. 4B**).

**Figure 4.**
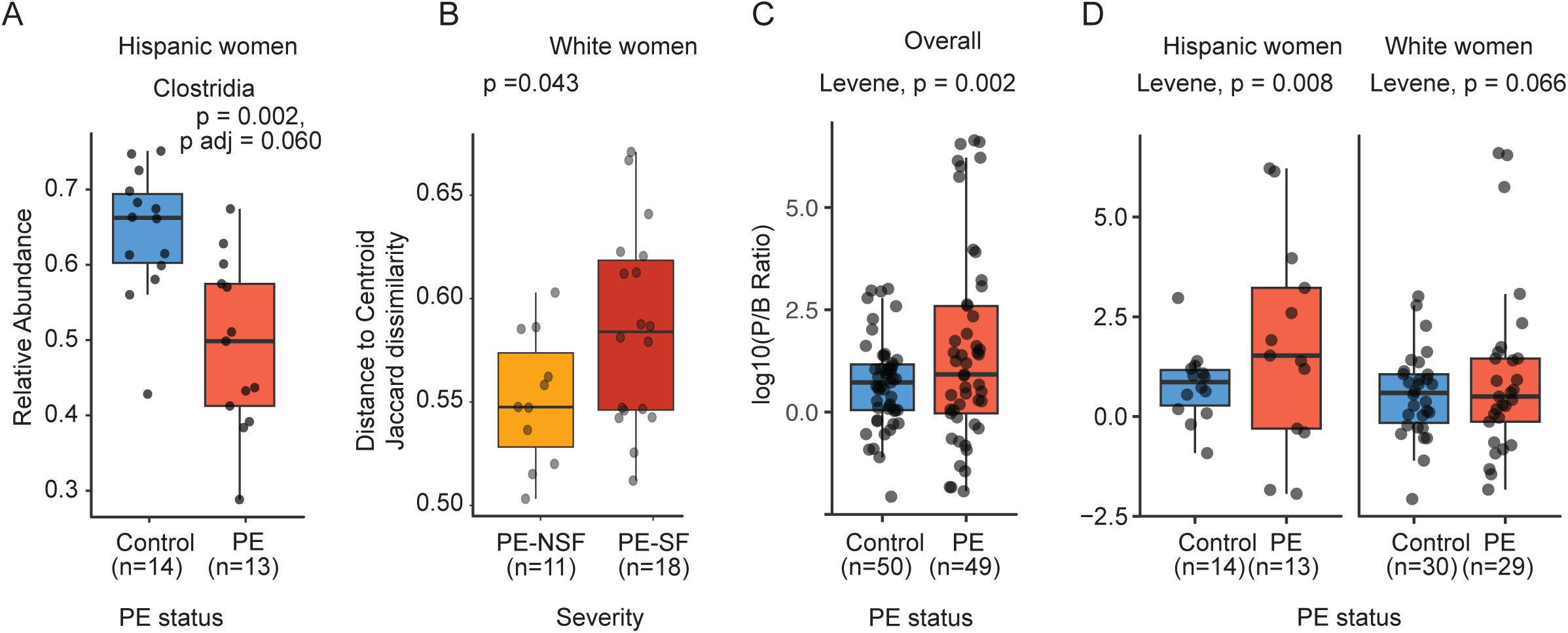
Women stool microbiome diversity in pregnant women according to preeclampsia (PE) status and severity, stratified by ethnicity (Hispanic or White). (A) Relative abundance of Clostridia class among Hispanic women according to PE status. (B) Beta dispersion analysis based on Jaccard distance among White women, comparing distance-to-centroid by preeclampsia severity. PE-SF, PE with severe features; PE-NSF, PE without severe features. (C, D) Variance of the log-transformed *Prevotella:Bacteroides* (P:B) ratio by preeclampsia status using Levene’s test in the overall population (C) and stratified by ethnicity (D).

To further explore the differences in microbial composition, we analyzed the *Prevotella:Bacteroides* (P:B) ratio in the stool samples. The P:B ratio reflects gut enterotypes and is shaped by diet, metabolism, and immune tone^48^; lower P:B ratios have been associated with pregnancy conditions, such as gestational diabetes mellitus^49^. We found no significant differences in the median P:B ratios by preeclampsia status or severity (**Table S9**). However, the variance in the P:B ratio was higher among women with preeclampsia overall (p = 0.0002) and among Hispanic women (p = 0.008), but not among White women (p = 0.066). No differences were observed by severity of preeclampsia (**Fig. 4C and D, and Table S9**).

In summary, while gut microbiome diversity did not differ by preeclampsia status or severity, women with preeclampsia showed a reduced prevalence of core taxa, such as *Alistipes* and *Bacteroides*, and an increased prevalence of *Facklamia_A*. Ethnicity-stratified analyses revealed decreased abundance and prevalence of Clostridia and lower prevalence of *Akkermansia* among Hispanic women with preeclampsia and increased interindividual variability among White women with severe features. Antibiotic use during pregnancy was associated with gut microbial diversity; however, ethnicity was not. While median P:B ratios did not differ by preeclampsia status, the variability in this ratio was greater among women with preeclampsia, particularly in the Hispanic group.

### Microbial diversity during delivery and blood pressure postpartum

Postpartum blood pressure measurements of this cohort were obtained 6 weeks postpartum during a standard visit. Both systolic and diastolic pressures at 6 weeks postpartum were significantly higher in women with preeclampsia than in the controls (**Fig. S3A**). When stratified by severity, diastolic pressure remained significantly higher in women with severe features than in those without, whereas systolic pressure did not differ between the two preeclampsia subgroups (**Fig. S3B**).

We then determined whether microbial diversity across maternal body sites at delivery was associated with postpartum blood pressure. Overall, only the oral microbiome demonstrated a significant association: women with higher oral alpha diversity had lower systolic and diastolic measures regardless of preeclampsia diagnosis (**Fig. S3C**). When stratifying by ethnicity, this association was significant only in Hispanic women (**Fig. S3D**) and was not observed in White women (**Fig. S3E**).

In the oral microbiota, lower postpartum diastolic blood pressure was associated with decreased abundance of Lachnospiraceae *G.2 sp.* (oral taxon 096) and *Leptotrichia sp.* (oral taxon 221) at delivery. Lower postpartum systolic blood pressure was associated with reduced abundance of *Prevotella sp.* (oral taxon 313), *Leptotrichia sp.* (oral taxa 221 and 417), and *Stomatobaculum longum* (**Fig. S4A and B**) at delivery. Among Hispanic women, we found negative associations between postpartum systolic blood pressure and *Prevotella histicola* and *Prevotella* sp. (taxon 313) (**Fig. S4C**) at delivery.

Regardless of preeclampsia diagnosis, higher oral microbial alpha diversity and lower abundance of specific taxa at delivery were associated with reduced blood pressure among Hispanic women at six weeks postpartum. These associations were not observed in the vaginal or gut microbiota.

### Infant microbiome

Of the 100 infants in this cohort, we successfully sequenced the stool microbiome of 79 infants: 39 born to women with preeclampsia and 40 born to controls. Sequencing success was associated with the timing of stool collection: samples collected closer to birth (mean = 0.43 days postpartum) were more likely to fail than those collected later (mean = 0.90 days postpartum), which were successfully sequenced. Infants with successful sequencing were demographically and clinically similar to the full cohort (**Tables 1 and S10**). Within this subset, however, infants born to mothers with severe features of preeclampsia had lower birth weights than those born to mothers without severe features, beyond the differences explained by gestational age alone (**Table S11**).

The infant gut microbial alpha diversity did not differ by maternal preeclampsia status (**Table S2**), but infants born to mothers with preeclampsia exhibited higher microbial interindividual variability for rare taxa than those born to mothers in the control group (p = 0.012, **Fig. 5A, Table S3**). Moreover, infants born to mothers with preeclampsia exhibited a lower prevalence of core taxa *Haemophilus_A* (Gammaproteobacteria class, 28.2% vs. 67.5%, p = 0.001, adjusted p = 0.061) and *YIM-78166* (Bacilli class, 41.1% vs. 72.5%, p = 0.006, adjusted p = 0.205) and a higher prevalence of core taxa *KA00134* (Clostridia class, 64.1% vs. 32.5%, p = 0.007, adjusted p = 205) than those born to controls (**Table S3**).

**Figure 5.**
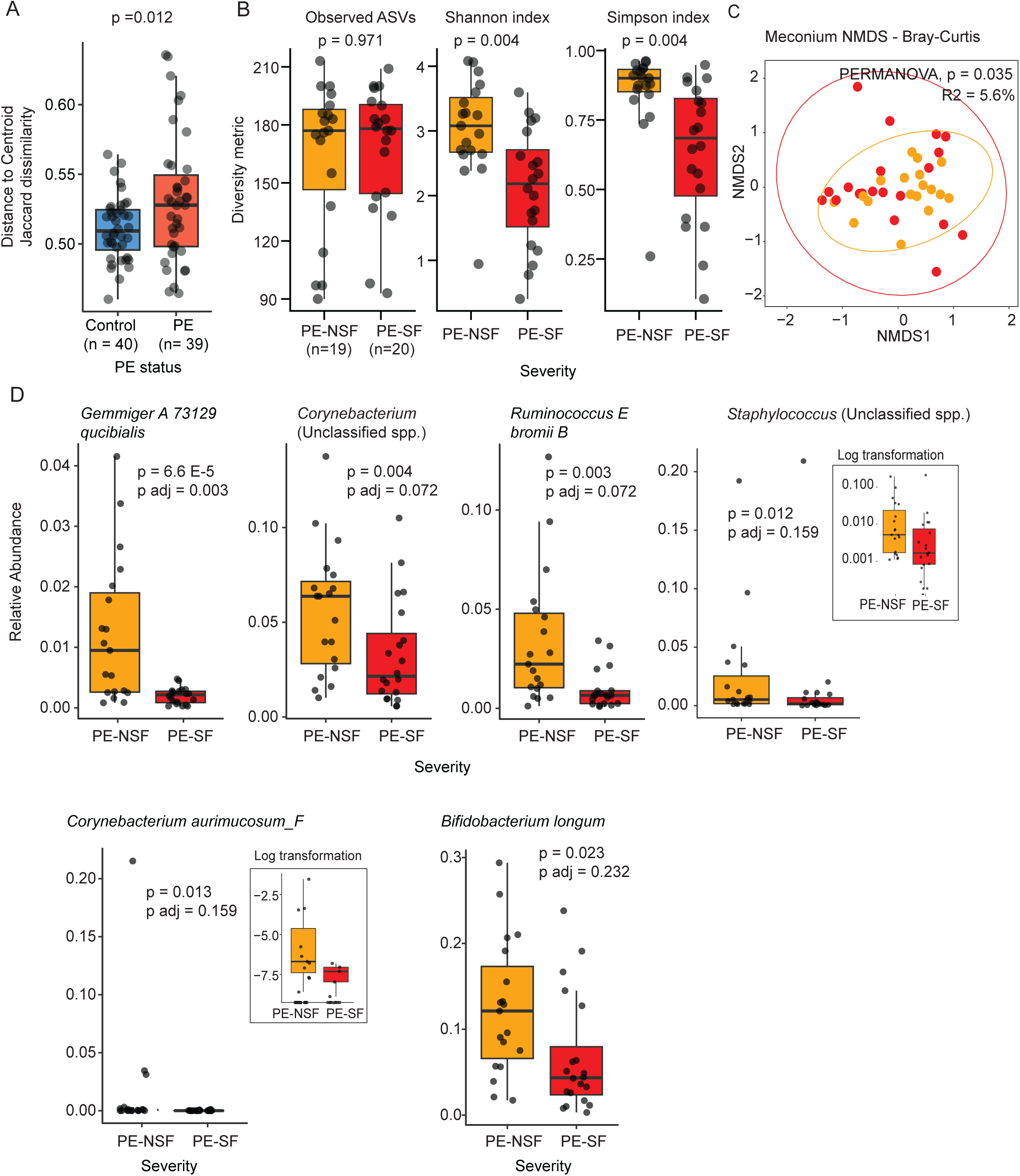
Infant stool microbiome diversity according to the mother’s preeclampsia (PE) status and severity. (A) Beta dispersion analysis based on Bray-Curtis distance, comparing distance-to-centroid between PE status. (B) Alpha diversity comparisons (Observed ASVs, Shannon, and Simpson indices) by PE severity. PE-SF, PE with severe features; PE-NSF, PE without severe features.; P values for alpha diversity analyses were obtained from linear models adjusting by BMI, antibiotic used during pregnancy and ethnicity. (C) Beta diversity analysis using Bray-Curtis distance and non-metric multidimensional scaling (NMDS) ordination; significance and R^2^ were assessed by PERMANOVA by to PE severity. Ellipses represent 95% confidence intervals. (D) Differentially abundant genera according to preeclampsia severity (p < 0.050, adjusted p < 0.250, MaAsLin2). The inset boxes show the log-transformed data.

Infants born to mothers with severe features of preeclampsia had significantly lower gut microbial alpha diversity than those born to mothers without severe features of preeclampsia or controls (p = 0.004; **Fig. 5B**). Notably, infants born to women without severe preeclampsia showed a gut microbial diversity similar to that of infants born to controls (**Fig. S2A**). When comparing the microbiome composition, there were no differences between the three groups (**Fig. S2B**), except when comparing only the preeclampsia groups by severity. The gut microbiome composition of the infants born to mothers with severe features of preeclampsia differed from those without severe features (p = 0.035, **Fig. 5C**). Moreover, infants born to mothers with severe features of preeclampsia exhibited decreased abundance of *Bifidobacterium longum, Corynebacterium* spp., *Ruminococcus_E. bromii_B*, *Corynebacterium aurimucosum_F*, and *Gemmiger_A_73129 qucibialis* (p < 0.050, adjusted p <;0.250, **Fig. 5D**).

In addition to preeclampsia severity, birth weight (which is closely associated with gestational age, but better explained diversity) and timing of sample collection were the strongest predictors of gut microbiome diversity (p < 0.050, **Table S2**); both variables were included as confounders in the models assessing microbial associations with preeclampsia.

In summary, we successfully sequenced the stool microbiome of 79 out of 100 infants in the cohort, with sequencing success associated with later sample collection times. While infant gut microbial diversity did not differ by maternal preeclampsia status, infants born to mothers with preeclampsia exhibited greater interindividual variability and distinct differences in the prevalence of specific core taxa. Infants born to mothers with severe preeclampsia had significantly lower microbial diversity and altered gut composition, including reduced abundance of *Bifidobacterium longum* and other commensals, than those born to mothers without severe features or to controls.

## Discussion

Preeclampsia remains a leading cause of maternal and perinatal morbidity globally^1,2^, with mounting evidence pointing to the maternal microbiome as a potential predictor of disease risk and contributor to pathogenesis. While prior studies have identified associations between preeclampsia and maternal microbiome, they have been limited by small sample sizes, a focus on a single body site, and a lack of ethnic diversity, with most being conducted outside the US^50^. In contrast, our study is the first to comprehensively profile the maternal microbiome across three distinct sites—oral, vaginal, and gut—in a large and ethnically diverse US cohort. In our study, the influence of preeclampsia on the maternal microbiome varied by ethnicity and body site, with the oral microbiome showing the strongest association. In contrast, vaginal and gut differences were subtler and mainly apparent after stratifying by ethnicity.

The oral microbiome of women with preeclampsia has been previously studied. Conflicting results regarding nitrate-reducing bacteria, namely, *Veillonella* abundance, reflect the complexity of microbiome research and the influence of multiple factors, including population characteristics, sampling methods, and study design^5,6,51,52^. In our patient cohort, women with preeclampsia exhibited a depletion of *Rothia,* while *Neisseria* levels were increased. *Rothia* and *Nesseria* are the primary nitrate-reducing bacterial genera in the oral cavity and are consistently abundant in individuals without oral diseases^53–55^. Among Hispanic women with preeclampsia, *Granulicatella adiacens* and *Haemophilus parainfluenzae* were depleted, while in White women, the key changes were *Rothia* loss and *Neisseria* increase, as observed for the overall population. Hispanic women lost taxa that, like *Rothia*, divert nitrite to ammonium through dissimilatory nitrate reduction to ammonium (DNRA)-like pathways. Both routes, however, converge on reduced oral NO bioavailability and impaired vascular regulation.

The decreased abundance of *Rothia* and increased abundance of *Neisseria,* specifically, *Neisseria subflava,* has also been reported previously^6^, and reflects the complex interplay between bacterial stress response capabilities and the inflammatory and oxidative stress environments characteristic of preeclampsia. For instance, *Rothia* species are vulnerable to the inflammatory and oxidative stress of preeclampsia because their iron-dependent metabolism is easily disrupted^56–59^. Their reliance on nitrate and lactate metabolism may not be sustained in an inflammatory environment, reducing their survival and competitiveness^60^. Conversely, *Neisseria* species adapt well to the inflammatory and oxidative stress of preeclampsia through strong defenses such as antioxidant enzymes, DNA repair, and stress response pathways^61–64^. Their ability to survive within activated neutrophils^65,66^ further confers a survival advantage in the heightened immune environment characteristic of preeclampsia. The metabolic fates of nitrite produced by *Rothia* and *Neisseria* differ. Although *Rothia* species possess nitrite reductase genes that can produce nitric oxide (NO) as an intermediate, the predominant pathway of nitrate utilization, DNRA, leads to ammonium formation rather than NO accumulation^53,67^. On the other hand, *Neisseria* predominantly utilizes the denitrification pathway to produce NO^68,69^. This distinction has profound implications for vasodilation. *Neisseria* species contribute directly to NO-mediated vasodilation through their denitrification pathway, whereas Rothia species, despite their superior nitrate reduction capacity, primarily produce ammonium rather than vasodilatory NO.

In the context of preeclampsia, the reduction of *Rothia* may represent a loss of nitrate-reducing capacity, and although the increase in *Neisseria* could act as a compensatory mechanism, it is unlikely to fully restore nitric oxide (NO) production under inflammatory conditions^5,70–73^. Overall, our findings on the oral microbiome align with studies demonstrating that nitrate-reducing bacteria may play a crucial role in blood pressure regulation through the nitrate-nitrite-nitric oxide pathway^5,51^, yet add an important aspect of strain specificity depending on ethnicity. Multiple studies have shown reduced nitrite levels in preeclamptic women^74,75^, supporting the concept that compromised nitric oxide synthesis, potentially mediated by oral microbiome alterations, is a key pathophysiological mechanism.

A decrease in the vaginal *Lactobacillus* abundance and increased microbial richness is linked to preeclampsia onset^6,7,76^. In this cohort, there were no differences in vaginal microbiome diversity according to preeclampsia status; however, women with severe features of preeclampsia showed a decreased abundance of *Lactobacillus jensenii* (Hispanics). The high prevalence of *Enterococcus* (White), *Fusobacterium*, and *Propionibacterium* in women with preeclampsia indicates a vaginal microbiome composition with a higher frequency of anaerobic, pro-inflammatory taxa compared to controls. We also observed that differences in vaginal microbiome diversity metrics among women with preeclampsia were mostly evident when analyses were stratified by ethnicity. Without this stratification, vaginal microbiome diversity appears largely similar between women with and without preeclampsia, underscoring ethnicity as a critical determinant of vaginal microbial composition and the necessity of including it as a central variable in both research and clinical approaches to preeclampsia^77–79^. The influence of ethnicity on the vaginal microbiome is well-established and multifactorial. For instance, genetic factors shape the vaginal environment and immune responses, favoring specific microbial communities in different populations^80–82^. Similarly, dietary patterns, which often correlate with cultural and ethnic backgrounds, impact nutrient availability and vaginal glycogen levels, affecting *Lactobacillus* dominance and microbiome stability^83–85^. Moreover, higher-quality diets support a more beneficial, *Lactobacillus*-dominated microbial community^83–86^. Finally, hygiene and sexual practices vary by culture and can disrupt or support a *Lactobacillus*-dominated environment^87,88^. These cultural factors interact to create unique vaginal microbial profiles that could account for differences in their association with preeclampsia.

Interestingly, Hispanic women with preeclampsia and severe preeclampsia exhibited reduced vaginal microbial diversity, which could reflect a compensatory response to microbial or vascular stress. However, larger sample sizes and characterization of the local inflammatory environment are needed. Whether reduced diversity is a driver or a consequence of preeclampsia remains uncertain.

The role of the gut microbiome in preeclampsia has also been studied in small cohorts outside the US, and the results show a reduced abundance of beneficial taxa and an increased presence of opportunistic pathogens in the condition (reviewed in^89^). However, there is considerable heterogeneity in the published findings, likely due to differences in study populations, methodologies, and environmental exposures. Contrary to other studies, we did not find differences in the gut microbiome of women according to preeclampsia status or severity.

Nonetheless, the reduced prevalence of Clostridia species in the gut, particularly among Hispanic women with preeclampsia, is relevant given the established role of Clostridia in Treg induction and activation^90–93^. Treg depletion is central to preeclampsia pathogenesis^1311,12^. Clostridia-derived SCFAs, particularly butyrate and acetate, directly promote Treg cell differentiation and expansion. Studies in mice have provided mechanistic evidence that a lack of SCFA bacteria can drive preeclampsia-like phenotypes through depletion of Tregs^11,12,94^. The decreased abundance of Clostridia class observed in Hispanic women with preeclampsia in this study may therefore contribute to reduced Treg populations, creating a pro-inflammatory environment that facilitates preeclampsia development. Similarly, our findings regarding *Akkermansia* prevalence are consistent with recent research demonstrating its protective role in preeclampsia^95,96^. In mice, treatment with *Akkermansia* enhanced gut barrier function, reduced endotoxemia, and promoted placental angiogenesis by restoring endothelial nitric oxide synthase (eNOS) activity and NO production.

The infant gut microbiome is seeded by maternal and environmental exposures, with early colonization patterns linked to future health outcomes. Previous studies have produced mixed results regarding the impact of maternal preeclampsia on the infant microbiome, with some suggesting that differences become more pronounced months after birth, as infants are continuously exposed to altered maternal microbiota^97,98^. The current findings show that immediate postnatal differences may be subtle, with differences in the prevalence of environmental or environment-sensitive bacteria (i.e., *Haemophilus_A*, *YIM-78166*, *KA00134*) among infants born to mothers with or without preeclampsia. Importantly, infants born to women with severe features of preeclampsia exhibited reduced abundance of *Bifidobacterium longum,* a beneficial member of the infant gut microbiome with known immunomodulatory function^99–101^.

Our study integrates multi-omics data, aligning with cutting-edge research efforts aimed at unraveling the complex system-level interactions that contribute to preeclampsia. Yet, several limitations should be acknowledged. Despite being the largest study of its kind in the US, the sample size of the preeclampsia cohort remains modest, limiting the statistical power, particularly for analyses adjusting for potential confounders. Additionally, stratification by ethnicity further reduced the analytical power, and groups with low representation had to be excluded from the subgroup analyses. Our approach was restricted to taxonomic profiling and did not assess microbial functional potential, which may be critical for understanding the mechanisms of host-microbe interactions. As a single-center observational study, we cannot infer causality or fully control for unmeasured environmental factors, such as diet, stress, or subclinical infections, that may have influenced the microbiome. Moreover, all samples were collected at the time of delivery, preventing the assessment of microbiome dynamics across pregnancy. These limitations highlight the need for larger, multi-ethnic, longitudinal studies that incorporate multi-omic and environmental data to identify predictive microbiome signatures and targets for early intervention. Despite these constraints, our findings contribute to the growing evidence linking the maternal microbiome to preeclampsia and support the rationale for future mechanistic and translational studies.

## Data Availability

The raw sequences and their associated metadata have been deposited in PRJNA1306676 (https://dataview.ncbi.nlm.nih.gov/object/PRJNA1306676?reviewer=e80pa9pcmannrthu27lv2tbeon). The complete code used for the bioinformatics and statistical analysis presented in this study is publicly available in the following GitHub repository: https://github.com/UMassDVR/Preeclampsia_microbiome_analyses_scripts.git

## Acknowledgments

We thank the pregnant women and infants who participated, UMass Memorial Health staff for recruitment and sample collection. We appreciate our colleagues’ feedback.

## Sources of Funding

This work was funded by a Novice Researcher Grant awarded to Dr. Courtney Birchall by The Gerber Foundation. The Gerber Foundation did not play a role in study design, analysis planning, or manuscript development.

## Disclosures

No disclosure.

## Notes

### Competing Interest Statement

The authors have declared no competing interest.

### Clinical Trial

This was not a clinical trial but an observational study

### Author Declarations

UMass Chan Institutional Review Board (Docket # H00020140)

